# When Second Best Might be the Best: Using Hospitalization Data to Monitor the Novel Coronavirus Pandemic

**DOI:** 10.1101/2020.05.11.20098475

**Authors:** Peter J. Mallow, Michael Jones

## Abstract

The novel coronavirus’ high rate of asymptomatic transmission combined with a lack of testing kits call for a different approach to monitor its spread and severity. We proposed the use of hospitalizations and hospital utilization data to monitor the spread and severity. A proposed threshold of a declining 7-day moving average over a 14-day period, “7&14” was set to communicate when a wave of the novel coronavirus may have passed. The state of Ohio was chosen to illustrate this threshold. While not the ideal solution for monitoring the spread of the epidemic, the proposed approach is an easy to implement framework accounting for limitations of the data inherent in the current epidemic. Hospital administrators and policy makers may benefit from incorporating this approach into their decision making.

## Introduction

Before government officials relax stay-at-home orders and hospitals resume elective procedures, decision-makers must accurately estimate the trend, severity, and prevalence of the novel coronavirus in a geographic region. Ideally, public health agencies would conduct active surveillance of infections in the general population (Sun, 2020; Nsubuga, 2006; Hashimoto, 2000). The results from this first-best solution represent a coincident indicator of COVID-19’s prevalence in a population. However, the fact that the novel coronavirus has a high rate of asymptomatic transmission hinders the usefulness of this approach (Gandhi, 2020).

Further hindering the disease surveillance is the limited number of novel coronavirus test kits as of April, 2020. Many states like Ohio prioritize the individuals who are eligible for testing (ODH, 2020a). Ohio and many other states recommend that all individuals who exhibit symptoms should be tested. However, hospitalized individuals and healthcare workers are given first priority. Individuals in long-term care and first responders are given a lower priority, and individuals in the general population have the lowest priority. While this prioritization redirects resources to their most effective use, the number of positive cases represents a biased sample of the general population. This tradeoff suggests that the number of positive test cases in a population does not necessarily reflect the actual prevalence of COVID-19 nor the infection rate trend.

As a lagging indicator, COVID-19 hospitalizations would normally be considered a second-best solution to measuring a trend in the infection rate. However, given the sample bias reflected in prioritized testing and asymptomatic transmission, we propose that COVID-19 hospitalizations combined with a capacity measure offer the best approach to measuring trends in COVID-19 infections. COVID-19 deaths present an even longer lag time than hospitalizations, and so they are not viewed as suitable of a measure. We chose the state of Ohio to illustrate our approach.

## Methods

The state of Ohio is one of several states that releases daily hospitalization data (ODH, 2020b). However they do not release length of stay (LOS) data. A literature search was performed in PubMed and the CDC Coronavirus website to identify studies published in March and April 2020 for LOS (CDC, 2020). A patient weighted pooled analysis was conducted to estimate the median LOS. The historical occupancy rate was obtained from the Centers for Disease Control and Prevention (CDC) National Center for Health Statistics for 2016 (CDC, 2016). Hospital capacity was defined as the number of staffed hospital beds (Definitive Healthcare, 2020).

The number of hospitalizations on a daily basis were multiplied by the median LOS to approximate the total number of bed days. Discharges based on LOS were subtracted to estimate a daily number of hospitalized COVID-19 patients. The number of occupied beds was calculated by multiplying the number of staffed beds by the pre-coronavirus occupancy rate. A 7-day moving average was calculated by adding the number of hospitalized COVID-19 patients over each seven-day window and dividing by the time period. The threshold for assessing the passing of a novel coronavirus wave was set at a declining 7-day moving average over a 14-day period. The moving average period of 7-days was chosen to mitigate daily and weekend reporting effects and to be consistent with prior epidemiologic models (Ng, 2020; Buckingham-Jeffery 2017; Rothman, 2008). The length of time was chosen based on the current knowledge of the high end of the novel coronavirus incubation period (Lauer, 2020). A further check included in the framework is stipulation that the 7-day moving average plus the historical occupancy level did not exceed the number of staffed beds during this window. The research was conducted with de-identified publicly available data and is exempt from institutional review board review. All analysis was conducted in Microsoft Excel (Microsoft, Inc. Redmond, WA).

## Results

The application of this approach to the state of Ohio found the first wave of the novel coronavirus passed on April 21, 2020. (Figure 1) During the period of January 7 to May 11, 2020, there were 4,413 COVID-19 hospitalizations. (Figure 2) Based on the median LOS of 4.9 days, these hospitalizations accounted for 21,624 hospital bed days (Table 1). The peak bed utilization based on the 7-day moving average occurred on April 7 with 4,340 COVID-19. At the peak, COVID-19 patients occupied 13.5% of the total staffed beds in Ohio. Combined with the occupancy rate, approximately 73% (23,979) of staffed beds would have been in use on the peak day, remaining under capacity. The results were based on an imputed LOS and occupancy level for Ohio and were intended to illustrate this approach rather than informing decision making.

**Figure 1.**
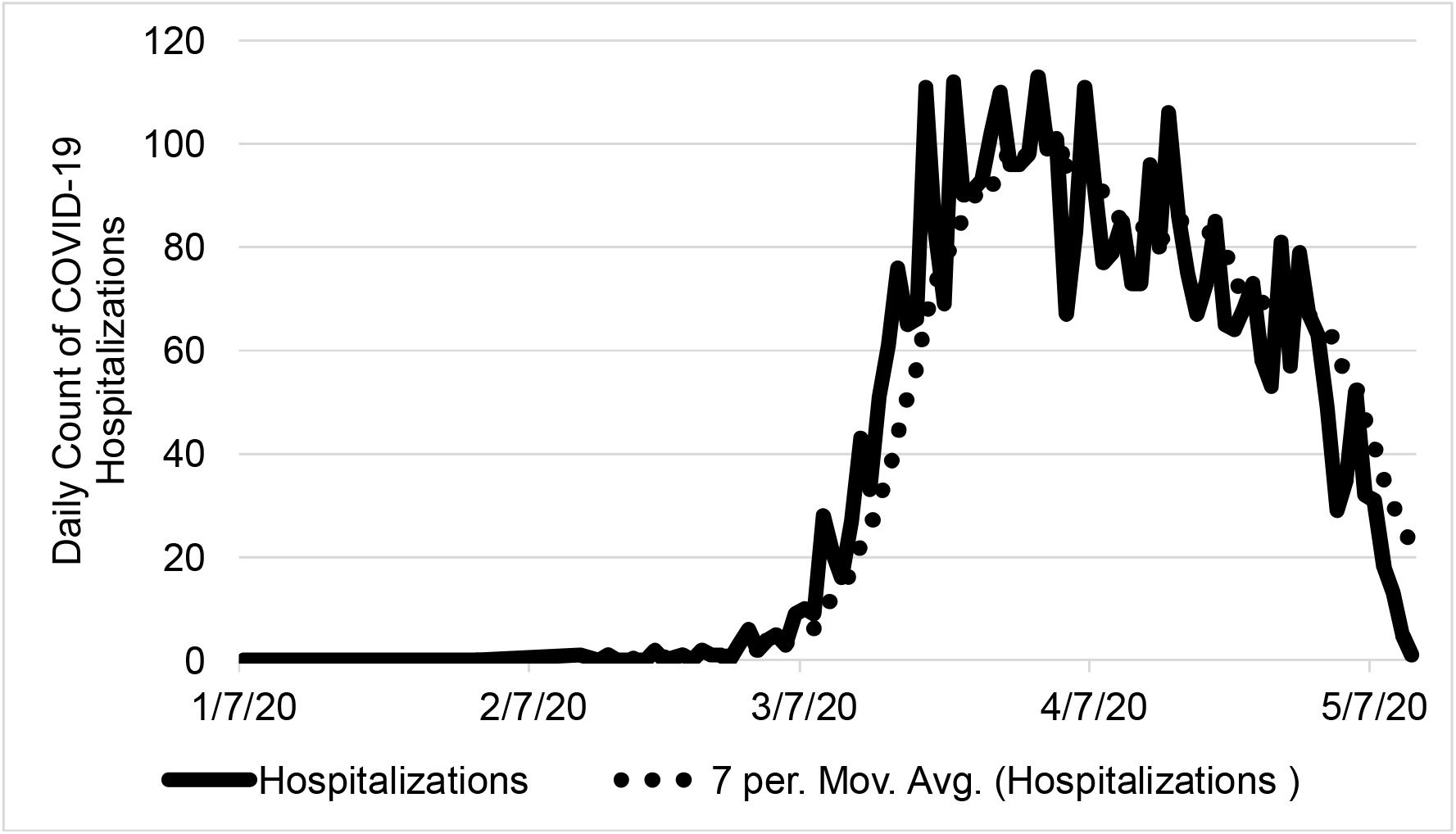
Ohio COVID-19 Occupied Hospital Beds by Day. The number COVID-19 occupied hospital beds is shown from January 7 to May 11, 2020 with the 7-day moving average.

**Figure 2.**
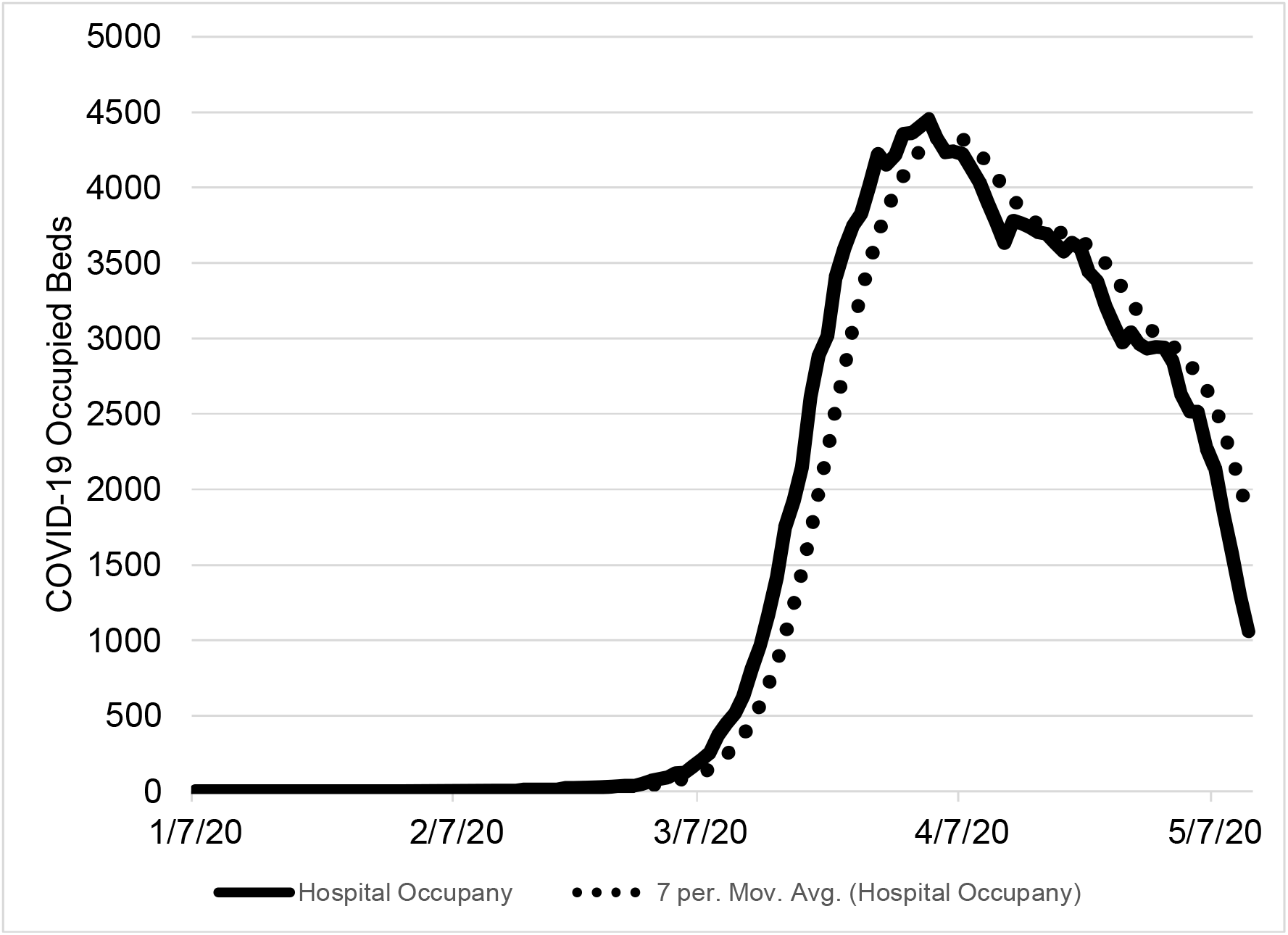
Ohio Daily COVID-19 Hospitalizations. The number daily COVID-19 hospitalizations are shown from January 7 to May 11, 2020 with the 7-day moving average.

**Table 1.**
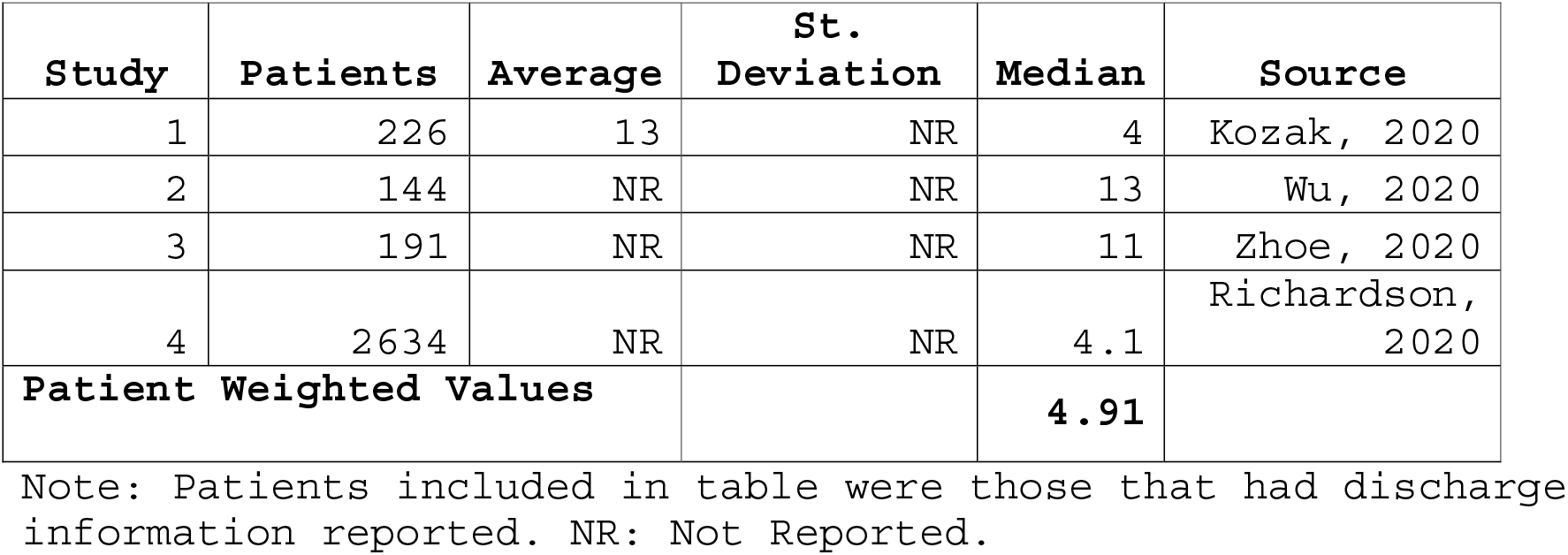
COVID-19 Hospital Length of Stay

| Study | Patients | Average | St.<br>Deviation | Median | Source |
| --- | --- | --- | --- | --- | --- |
| 1 | 226 | 13 | NR | 4 | Kozak, 2020 |
| 2 | 144 | NR | NR | 13 | Wu, 2020 |
| 3 | 191 | NR | NR | 11 | Zhoe, 2020 |
| 4 | 2634 | NR | NR | 4.1 | Richardson,<br>2020 |
| **Patient Weighted Values** |  |  |  | **4.91** |  |

Note: Patients included in table were those that had discharge information reported. NR: Not Reported.

## Discussion

A critical component of monitoring the novel coronavirus pandemic is availability of reliable and valid data. Preferably, we would have widespread testing data to inform our epidemiological models and provide a leading indicator of future demands of our healthcare system. However, the widespread asymptomatic community transmission and lack of testing kits prevents us from having a clear understanding of the novel coronavirus spread. In the absence of wide spread testing prior to or at the initial onset of the epidemic, hospitalizations and hospital utilization become the second-best indicator to monitor the severity and progression of the novel coronavirus. Hospital utilization must be monitored to ensure that the hospitalization raw numbers do not become truncated. Once hospitals approach maximum capacity, the hospital’s decision to triage and an individual’s decision to seek care elsewhere or stay at home will introduce bias into the data measure. This necessity to avoid a biased indicator was the motivating reason to track hospitalizations in the first place. In geographic regions that are approaching capacity or where hospitals are already at maximum utilization, hospitalizations may be less indicative of COVID-19’s prevalence. If this stage is reached however, any discussion about opening up hospitals for elective procedures is moot.

Using a novel data set from Ohio, this proposed framework provided a means to illustrate the monitoring and severity of the novel coronavirus while adjusting for daily fluctuations in the data. Our threshold of a declining 7-day moving average over a 14-day period, “7&14,” provided a conservative threshold for informing public policy decisions, such as access to healthcare services, regarding the novel coronavirus pandemic.

Our approach is broadly consistent with the work of the University of Minnesota (UM), Carlson School of Management (Health Affairs, 2020). The UM initiated a COVID-19 Hospitalization Tracking Project, and our work expands upon the efforts of UM by incorporating hospital capacity and providing a means to assess the ongoing epidemic. Baker et al. 2017 proposed an approach for tracking influenza intensive care unit bed utilization to monitor severity of the influenza season (Baker, 2017). However, many states are not reporting hospitalizations reliably or at all, let alone intensive care beds to provide usual information that can be aggregated.

The proposed “7&14” framework has two key advantages. First, it can be implemented at the individual hospital level and aggregated by geographic regions. It requires three data inputs, hospitalizations, LOS, and occupancy. Second, one of the inherent benefits of using a moving average is to smooth out random short-term fluctuations in daily hospitalizations. These two attributes combined creates an easy to understand dashboard at the chosen level of analysis to assess the severity and spread of the novel coronavirus epidemic. If or when additional healthcare system supply data becomes available (i.e. intensive care bed utilization), this approach can easily be expanded.

This proposed approach can inform public decisions regarding the resumption or curtailment of healthcare service and monitor the ascent or descent of waves of the novel coronavirus. It provides a means to monitor the situation on a daily basis as new data emerges and report in a clear and concise manner the current nature of the novel coronavirus epidemic.

The proposed “7&14” approach, leveraging hospitalizations and hospital utilization, may not be the ideal method of monitoring the novel coronavirus epidemic. However, widespread community asymptomatic transmission and lack of testing kits, elevates this approach to the best available. With improved reporting of COVID-19 hospitalizations, LOS, and hospital occupancy across the country this approach may improve decision making for hospital administrators and policy makers.

## Data Availability

The authors will make all data available upon reasonable request.

## Acknowledgements

The authors would like to thank Michael Topmiller, PhD, Edmond A. Hooker, MD, DrPH, Dee Ellingwood, MS, and Jennifer Mallow, MBA for their thoughtful comments and suggestions.

## Funding and Conflict of Interest

The authors did not receive any funding for this research nor have any conflicts of interest regarding this research to report.

## Author Contribution

All authors contributed equally to all aspects of this reseach and manuscript.

## Notes

### Competing Interest Statement

The authors have declared no competing interest.

